# Automatic quantification of brain lesion volume from post-trauma MR Images

**DOI:** 10.1101/2021.04.24.21255599

**Authors:** Thomas Mistral, Pauline Roca, Christophe Maggia, Alan Tucholka, Florence Forbes, Senan Doyle, Alexandre Krainik, Damien Galanaud, Emmanuelle Schmitt, Stéphane Kremer, Adrian Kastler, Irène Troprès, Emmanuel L Barbier, Jean-François Payen, Michel Dojat

**Affiliations:** Univ. Grenoble Alpes, Inserm, CHU Grenoble Alpes, GIN, 38000 Grenoble, France; Pixyl, 38000 Grenoble, France; Univ. Grenoble Alpes, Inria, CNRS, Grenoble INP, LJK, 38000 Grenoble, France; APHP, Hôpital Pitié Salpétrière, 75000 Paris, France; CHU, Hôpital Central, 54000 Nancy, France; CHU, de Strasbourg, F-67000 Strasbourg, France; Univ. Grenoble Alpes, Inserm, CHU Grenoble Alpes, CNRS, IRMaGe, 38000 Grenoble, France

**Author notes:** **Corresponding author:** Michel Dojat, Grenoble Institut des Neurosciences, INSERM Chemin F. Ferrini 38700 La Tronche, France, Ph : 33 (0)4 56 52 06 01. T. Mistral; P. Rocca; C. Maggia:; A. Tucholka; F. Forbes; S. Doyle; A. Krainik; D. Galanaud; E. Schmitt; S. Kremer; A. Kastler; I. Troprès; E. L. Barbier; JF Payen.

**Keywords:** MRI, Mean Diffusivity, Segmentation, Traumatic brain lesion

## Abstract

**Objectives:** The determination of the volume of brain lesions after trauma is challenging. Manual delineation is observer-dependent and time-consuming which inhibits the practice in clinical routine. We propose and evaluate an automated atlas-based quantification procedure (AQP) based on the detection of abnormal mean diffusivity (MD) values computed from diffusion-weighted MR images.

**Methods:** We measured the performance of AQP versus manual delineation consensus by independent raters in two series of experiments: i) realistic trauma phantoms (n=5) where abnormal MD values were assigned to healthy brain images according to the intensity, form and location of lesion observed in real TBI cases; ii) severe TBI patients (n=12 patients) who underwent MR imaging within 10 days after injury.

**Results:** In realistic trauma phantoms, no statistical difference in Dice similarity coefficient, precision and brain lesion volumes was found between AQP, the rater consensus and the ground truth lesion delineations. Similar findings were obtained when comparing AQP and manual annotations for TBI patients. The intra-class correlation coefficient between AQP and manual delineation was 0.70 in realistic phantoms and 0.92 in TBI patients. The volume of brain lesions detected in TBI patients was 59 ml (19-84 ml) (median; 25-75^th^ centiles).

**Conclusions:** our results indicate that an automatic quantification procedure could accurately determine with accuracy the volume of brain lesions after trauma. This presents an opportunity to support the individualized management of severe TBI patients.

**Key points:**

- The management of patients with severe traumatic brain injury is complex, and access to objective quantitative information lesion volumes can support clinical decision-making.
- An automated delineation procedure was developed to determine the nature and volume of brain lesions post-trauma.
- This procedure was based on diffusion weighted MR-imaging to quantify the volume of vasogenic and cellular edema from realistic phantoms and patients with severe traumatic brain injury.
- Nature and quantification of the brain lesions volume compared favorably with manual delineation of brain lesions by a panel of experts.

## Introduction

Traumatic Brain Injury (TBI) remains a leading cause of death and disability among young people. A small proportion of patients with severe TBI, as defined by an initial Glasgow Coma Scale (GCS) score of less than 9, will not have disabilities [1]. Predicting neurological outcome after severe TBI is challenging due to the complexity of the traumatic lesion, its evolution over time, and the number of external factors that may affect the outcome. Nevertheless, the determination of nature and volume of brain lesion has been identified as a clinically relevant criterion in estimating outcome [2].

Diffusion-weighted imaging (DWI) is a technique sensitive to detecting subtle microstructural changes in white matter tracts, and particularly suitable for identifying edema and necrosis [3; 4]. A reduction of mean diffusivity (MD) is related to cellular (cytotoxic) edema while an increase of MD indicates a vasogenic edema [5]. Both types of brain edema exist at the acute phase of severe TBI, and are major contributors to the elevation of intracranial pressure and poor outcome after TBI [6]. Although DWI compares favorably with clinical/radiographic prognosis scores [7], there is still insufficient evidence to recommend DWI as a prognostic method for TBI patients [8; 9]. This may be due to limited data using automated methods to quantify brain injury post-trauma [5; 7]. Skull deformation, intracranial blood in the brain tissue, the presence of cerebral spinal fluid (CSF) and the heterogeneity of brain tissue injury make the segmentation of traumatic brain lesion challenging. Automated approaches using non-contrast CT imaging were developed for cranial cavity segmentation[10], cistern segmentation or detection of intracranial hematomas [11]. Due to the higher sensitivity of MRI compared to CT scan [12], more intracranial lesions (e.g. brain swelling or intracranial hemorrhage) could be detected. For its ability to distinguish both types of brain edema, MD was chosen in the present study.

Our aim was to develop an automated approach to quantify post-traumatic edematous brain lesion volumes using MD values from DWI. A quality control procedure was implemented to account for the high dependence of DWI on scanning equipment and acquisition protocol [13; 14]. We constructed an automated atlas-based quantification procedure (AQP) to partition the brain into defined regions, and detect voxels with abnormal MD values, i.e. vasogenic and cellular, within those regions. The validation was performed using both realistic phantoms and severe TBI patients. We compared the automated delineation results to the manual delineations performed by expert raters.

## Methods

Two sets of experiments were performed (**Fig.1**): i) realistic TBI phantoms with artificially introduced lesions with abnormal MD values and ii) TBI patients who were part of the validation process of MRI acquisition from an ongoing multicenter clinical trial. The study was approved by the local ethical committee (Comité de Protection des Personnes Sud-Est V, ID RCB-2014-A01674-43) and registered with ClinicalTrial.gov (OxyTC, NCT02754063).

**Figure 1.**
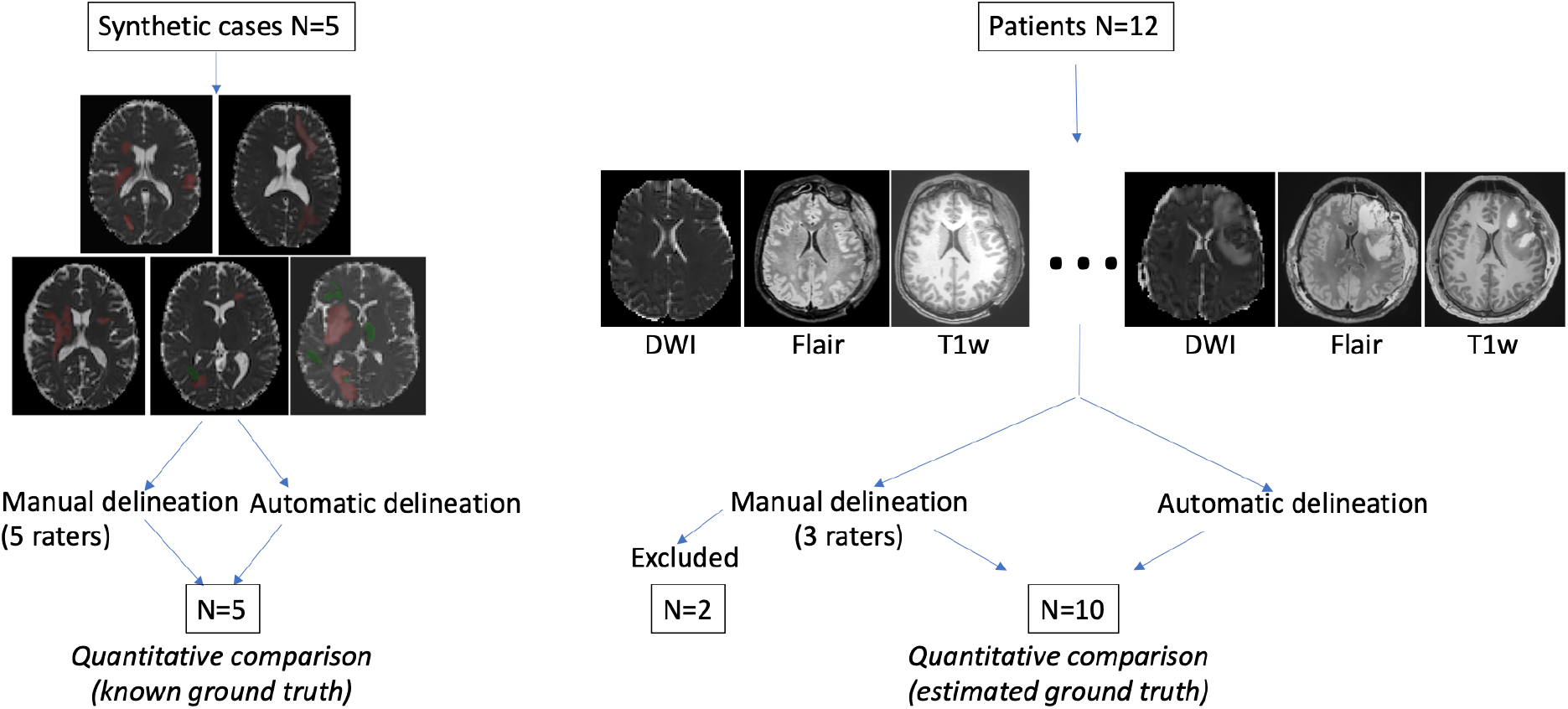
Evaluation procedure. Left: five realistic TBI lesion cases were constructed with low level (green) and high level (red) artificial values. By construction the ground truth was known for automatic and manual lesion delineation comparison. Right: Twelve TBI patients were included, each with three types of MR images. Manual and automatic delineation results were quantitatively compared for 10 patients. The ground truth was defined as the consensus of expert annotations, calculated using Staple.[16]

Manual delineation was performed by a panel of five expert neuroradiologists.

### Realistic TBI phantoms

DW imaging was performed on 5 healthy volunteers (Philips Achieva 3.0T TX, Philips Healthcare, Best, the Netherland) at the IRMaGe MRI facility (Grenoble, France). Low and high MD values, simulating cellular and vasogenic brain edema respectively, were manually inserted in these brain images by a neuroradiologist (TM) familiar with traumatic lesions. The simulated values were obtained by the application of a multiplicative coefficient to the real MD values. The range of the coefficient was 0.41-0.91 and 1.10-2.10 for low and high MD, respectively. A Gaussian filter (3 mm half-width) was applied in accordance with observed TBI edema appearance. The MD maps were modified exclusively; the corresponding anatomical images remained unmodified.

### TBI patients

One patient (Supplementary Material, SM: Table 1 for inclusion and non-inclusion criteria) from each of 12 participating sites underwent an MRI exam (SM: Table 3 for details) between 5 to 13 days after trauma. At each site, additional DWI images were acquired from 3 healthy volunteers (controls, see SM: Table 2 for inclusion and non-inclusion criteria), to compute reference MD maps. The images from each site were anonymized, uploaded and stored in a dedicated centralized academic imaging data repository (shanoir.irisa.fr).

**Table 1.**
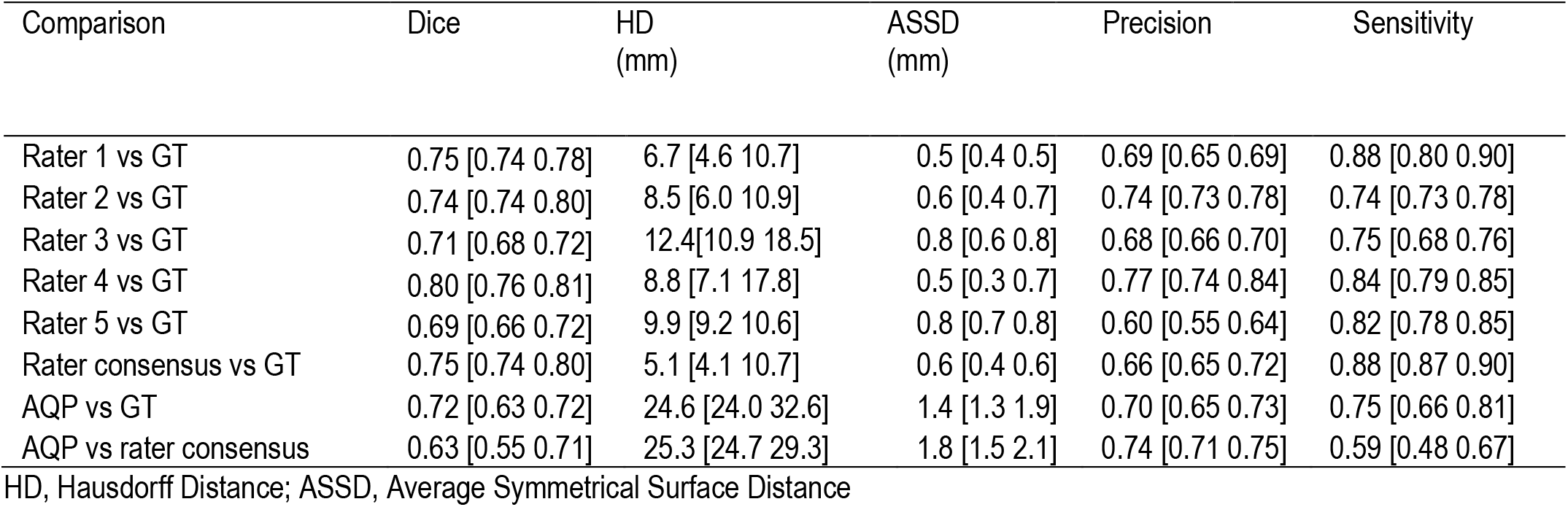
Spatial measures for the 5 realistic TBI phantoms. Each rater, rater consensus and automatic quantification procedure (AQP) were compared to the ground truth (GT) as reference. Data are expressed as median and 25-75^th^ percentiles. Dice and precision obtained from rater consensus and AQP were comparable. HD and ASSD were higher using AQP compared to rater consensus (P<0.05).

| Comparison | Dice | HD<br>(mm) | ASSD<br>(mm) | Precision | Sensitivity |
| --- | --- | --- | --- | --- | --- |
| Rater 1 vs GT | 0.75 [0.74 0.78] | 6.7 [4.6 10.7] | 0.5 [0.4 0.5] | 0.69 [0.65 0.69] | 0.88 [0.80 0.90] |
| Rater 2 vs GT | 0.74 [0.74 0.80] | 8.5 [6.0 10.9] | 0.6 [0.4 0.7] | 0.74 [0.73 0.78] | 0.74 [0.73 0.78] |
| Rater 3 vs GT | 0.71 [0.68 0.72] | 12.4 [10.9 18.5] | 0.8 [0.6 0.8] | 0.68 [0.66 0.70] | 0.75 [0.68 0.76] |
| Rater 4 vs GT | 0.80 [0.76 0.81] | 8.8 [7.1 17.8] | 0.5 [0.3 0.7] | 0.77 [0.74 0.84] | 0.84 [0.79 0.85] |
| Rater 5 vs GT | 0.69 [0.66 0.72] | 9.9 [9.2 10.6] | 0.8 [0.7 0.8] | 0.60 [0.55 0.64] | 0.82 [0.78 0.85] |
| Rater consensus vs GT | 0.75 [0.74 0.80] | 5.1 [4.1 10.7] | 0.6 [0.4 0.6] | 0.66 [0.65 0.72] | 0.88 [0.87 0.90] |
| AQP vs GT | 0.72 [0.63 0.72] | 24.6 [24.0 32.6] | 1.4 [1.3 1.9] | 0.70 [0.65 0.73] | 0.75 [0.66 0.81] |
| AQP vs rater consensus | 0.63 [0.55 0.71] | 25.3 [24.7 29.3] | 1.8 [1.5 2.1] | 0.74 [0.71 0.75] | 0.59 [0.48 0.67] |

HD, Hausdorff Distance; ASSD, Average Symmetrical Surface Distance

**Table 2.**
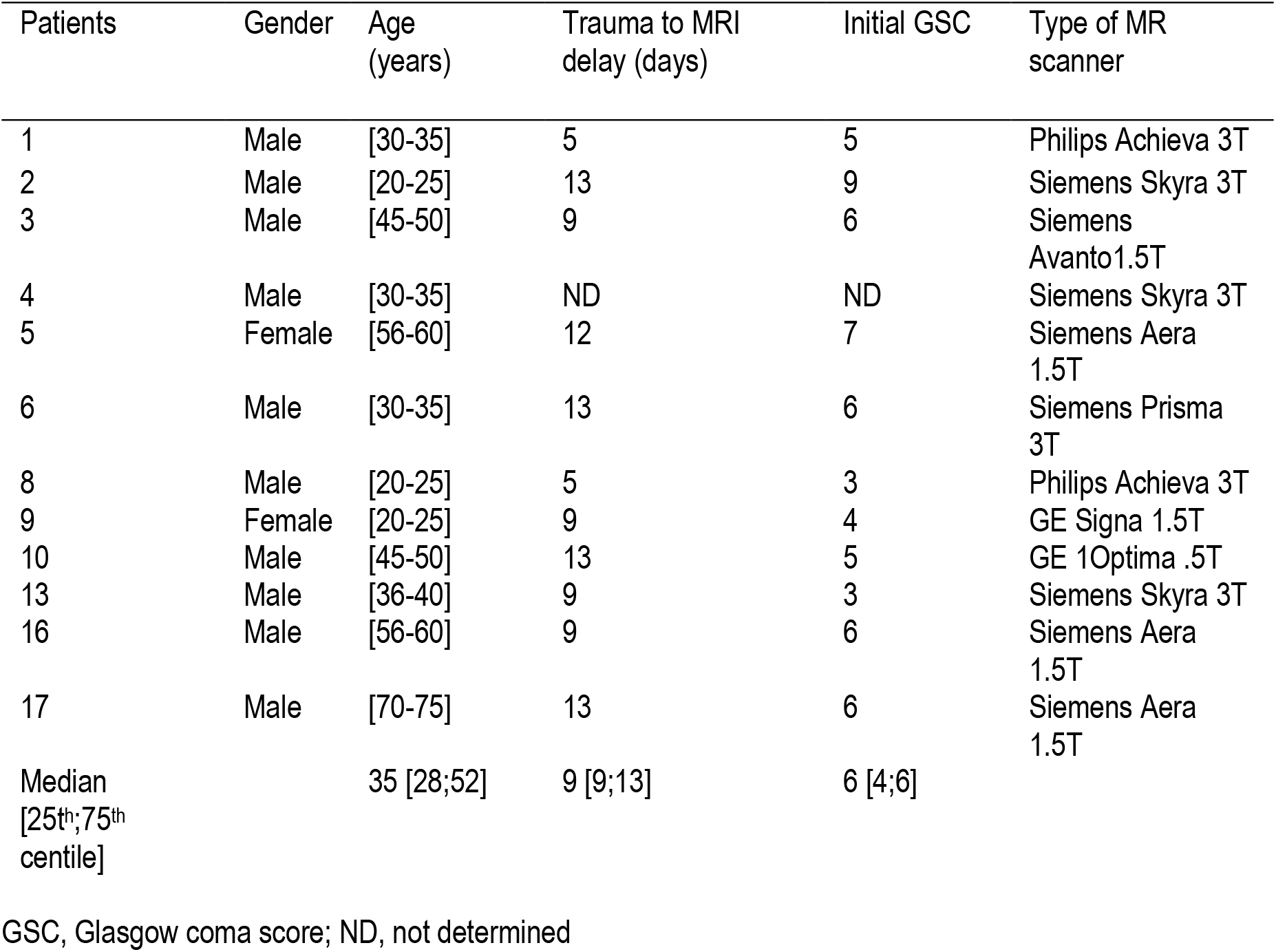
Characteristics of the 12 patients with severe traumatic brain injury. Patients #1 and #10 were excluded from the analysis because one rater delineated brain lesions on FLAIR images.

**Table 3.** Spatial measures for the 10 patients with severe traumatic brain injury. The automatic quantification procedure (AQP) is compared to the consensus from 3 raters. HD, Hausdorff Distance; ASSD, Average Symmetrical Surface Distance.

| Patients | Dice | HD<br>(mm) | ASSD<br>(mm) | Precision | Sensitivity | AQP: Vol.<br>edema (ml) | AQP: Vol.<br>vasogenic<br>edema (ml) | AQP: Vol.<br>cellular<br>edema (ml) |
| --- | --- | --- | --- | --- | --- | --- | --- | --- |
| 2 | 0.49 | 32.08 | 3.19 | 0.42 | 0.58 | 55.2 | 36.7 | 18.4 |
| 3 | 0.61 | 15.0 | 2.93 | 0.62 | 0.61 | 13.9 | 87.0 | 5.1.9 |
| 4 | 0.73 | 38.39 | 1.86 | 0.64 | 0.86 | 80.9 | 67.8 | 13.1 |
| 5 | 0.78 | 25.32 | 1.04 | 0.78 | 0.78 | 84.6 | 79.2 | 5.3 |
| 6 | 0.71 | 32.95 | 1.15 | 0.72 | 0.70 | 33.1 | 28.4 | 4.7 |
| 8 | 0.52 | 20.12 | 2.12 | 0.67 | 0.43 | 7.2 | 57.5 | 1.5 |
| 9 | 0.43 | 33.66 | 3.73 | 0.49 | 0.39 | 6.9 | 30.3 | 3.9 |
| 13 | 0.56 | 25.63 | 1.80 | 0.49 | 0.64 | 240.6 | 72.8 | 167.7 |
| 16 | 0.78 | 11.79 | 0.85 | 0.82 | 0.74 | 130.3 | 121.0 | 9.2 |
| 17 | 0.33 | 36.77 | 5.84 | 0.22 | 0.64 | 63.7 | 45.12 | 18.4 |
| Median | 0.58 | 28.8 | 2.0 | 0.63 | 0.64 | 59.4 | 41.0 | 7.3 |
| [25<sup>th</sup>;75<sup>th</sup><br>centile] | [0.50;0.72] | [21.4;33.5] | [1.3;3.1] | [0.49;0.70] | [0.59;0.73] | [19;84] | [14;72] | [5;17] |

### Quality Control Procedure

A quality control procedure was developed and deployed as the Pixyl research platform (**Fig. 2**). Automatic procedures analyzed specific DICOM tags, susceptibility artifacts, signal-to noise ratio, motion, and corrupted slices. A quality control report was provided and validated by MR physicists (IT, CM).

**Figure 2.**
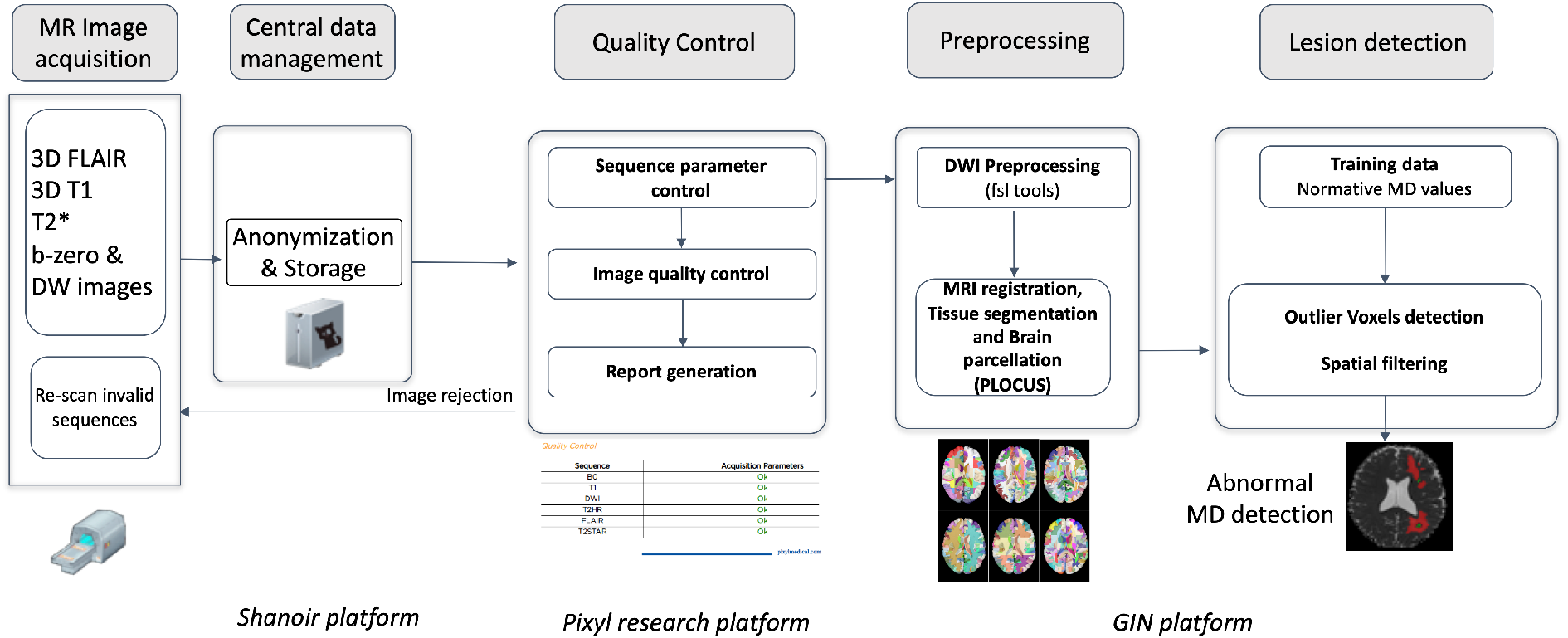
Image processing pipeline from image acquisition to mean diffusivity abnormalities automatic detection.

### Manual delineation

A panel of five expert neuroradiologists, more than 10 years’ experience (AKa, AKr, DG, ES and SK), manually annotated brain lesion areas from realistic TBI phantoms. The panel was not aware that the lesions were manually inserted.

Three of these experts (AKr, DG and SK) then manually annotated brain lesions from TBI patients. They followed an annotation protocol based on DWI and the ITK-SNAP tool (http://www.itk.org) for annotation, blinded to each other and the ground truth. Other MRI sequences could be used for additional cues. Because manual delineation has inherent inter-rater variability [15], the Simultaneous Truth and Performance Level Estimation (Staple) method was used to provide an estimation of the rater consensus [16].

### Automated Quantification Procedure (AQP)

Diffusion source images were denoised [17], corrected for inter-volume subject motion and geometric distortion (**Fig. 2** and SM); and MD maps were computed from the trace of the diffusion tensor (see SM: Table 3). Brain was extracted and segmented using a Bayesian Markov Random Field approach named PLOCUS [18].

AQP used 6 parcellation atlases to establish normative values and detect abnormal voxels according to the Potholes and Molehills method [19; 20]. A voxel was considered as abnormal if its values deviated outside the normal range in ≥4 parcellation atlases. Voxels exhibiting high and low MD were considered if they formed part of a lesion of minimum size 0.16ml and 0.12ml, respectively. Voxels from CSF or ventricles, as defined by segmentation of the T1-w sequence, were excluded. To deal with partial volume effects, abnormal high MD voxels at a distance less than 3 mm from CSF voxels were also excluded. Lesion volume was expressed in ml and in brain volume fraction (%) that reflects the ratio between brain lesion volume and supra-tentorial brain volume.

### Quantitative comparison of the manual and automatic delineation methods

Five spatial measures were used to compare delineation methods: the Dice metric to measure the volume overlap, the Average Symmetrical Surface Distance (ASSD) to measure the average Euclidian surface distance, the Hausdorff Distance (HD) to measure the maximum distance between two surface points, and Precision and Recall (sensitivity) to assess over- and under-segmentation, respectively (see http://www.isles-challenge.org/ISLES2015/ for formulas). For ASSD and HD, expressed in mm, optimal values tend to 0. For Dice, Precision and Recall values, expressed within a 0-1 range, optimal values tend to 1.

### Statistical analysis

Data were expressed as mean ± standard deviation or median (25-75^th^ centiles). The Intra-class Correlation Coefficient (ICC) was used to measure the reliability of measurements between the rater consensus and AQP. The nonparametric Kruskall-Wallis test was used to compare spatial measures between GT, AQP, the rater consensus, and each rater (realistic phantom). The Mann-Whitney test was used to compare the rater consensus and AQP (TBI patients). Statistical significance was established when P< 0.05.

## Results

### Realistic TBI phantoms

The mean volume of the lesions manually inserted was 31 ml, i.e. 2.2 % of the total brain volume, and corresponded to the ground truth (GT). Typical examples of agreements between GT, manual delineation, and AQP are shown in Fig.3. Compared to manual delineation, AQP could detect additional lesions that were present in GT and could exclude imaging artefacts. The time taken to process each case was 30 minutes for manual delineation versus 10 minutes using AQP.

Dice and precision obtained with manual delineation did not significantly differ from those with AQP (Dice: 0.75 and 0.72 and Precision: 0.66 and 0.70 respectively) (**Table 1**). The surface distance measures of HD and ASSD were significantly higher using AQP compared to manual delineation (both P<0.05).

The lesion volumes corresponded to 2-4% of the brain volume, i.e. 18-40 ml. Both raters and AQP overestimated the lesion volumes in realistic phantoms, compared to GT (+32% for rater consensus; +13% for AQP) (**Fig. 4**, SM: Table 4). Note that the raters have a more consistent relationship with ground truth though tending to overestimate, while AQP has some jitter. The reliability between rater consensus ratings and AQP was moderate (ICC = 0.70) (**Fig. 4** and SM: Table 4). Of note was the overestimation of brain lesion volume with high MD values by raters whilst low MD lesion volumes were underestimated (SM: Table 5 and Table 6). However, there was no difference between AQP, the rater consensus and GT regarding the determination of brain lesion volumes (P=0.27).

**Figure 4.**
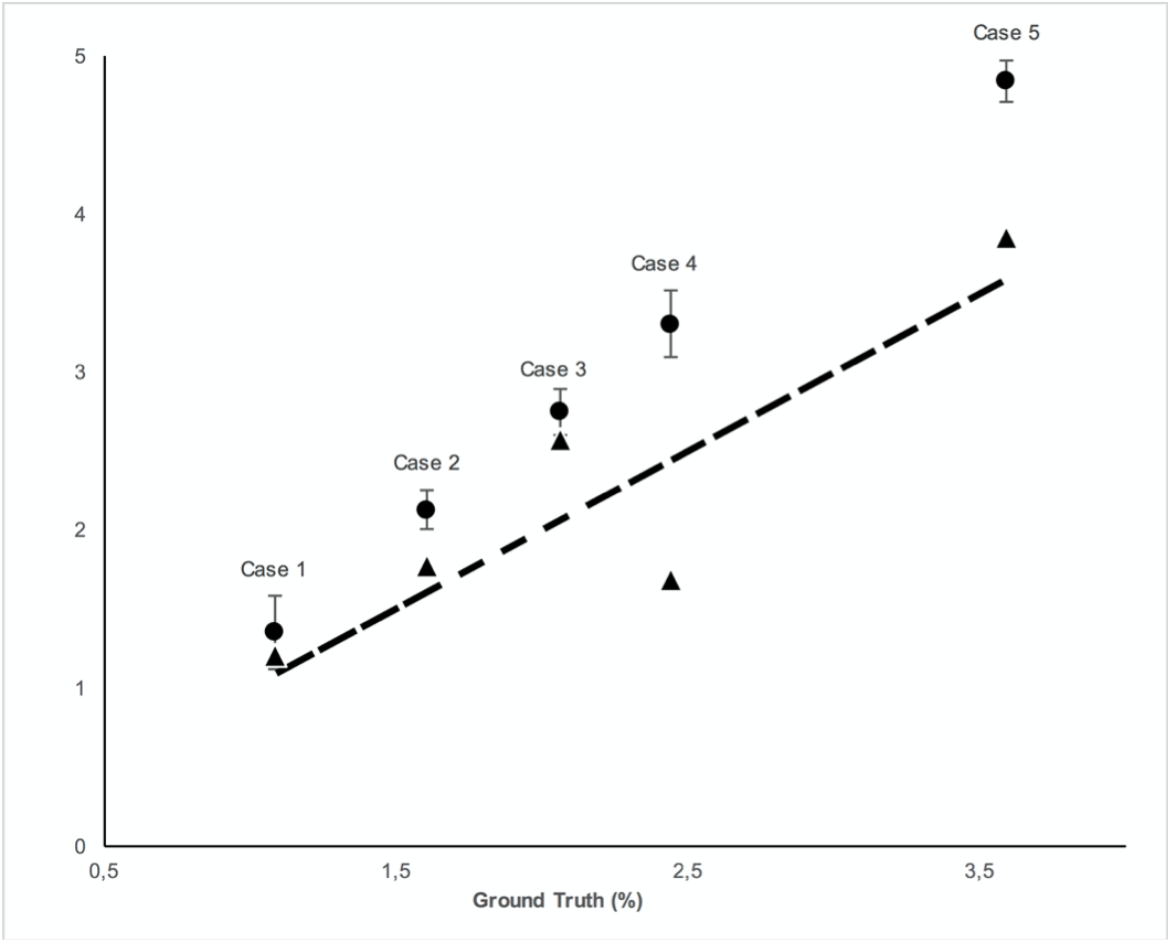
Total lesion volume (low + high MD) in % brain volume of diffusion-weighted images (DWI) for five healthy volunteers (realistic TBI phantoms). Lesion volumes (mean, 95% confidence interval) were determined using manual delineation by 5 raters (black circles) and automated quantification procedure (black triangles) (Y-axis) versus ground truth (X-axis). The dashed line indicates the identity curve.

### TBI patients

The characteristics of the patients are shown in **Table 2**. Two patients (#1 and #10) were excluded from the analysis because one rater delineated brain lesions visible on FLAIR images only. **Fig. 5** shows low and high MD brain lesions depicted by the rater consensus (middle) and by AQP (right). Additional brain lesions were found using AQP (cf. S2 and S17 in **Fig. 5**). Dice, precision and sensitivity were comparable between AQP and rater consensus (SM: Table 7). Surface distance measures of HD and ASSD were slightly different (median 28.8 and 2.0 mm for AQP vs 19.6 and 1.4 mm for raters, respectively with P<0.02 for the former, non-significant for the latter) (SM: Table 7). The brain lesion volumes of these patients computed by AQP ranged from 0.4% to 14.7% of the brain volume, i.e. 7 to 241 ml, including 41 ml (14-72 ml) (median; 25-75^th^ centiles) and 7 ml (5-17 ml) for high (vasogenic edema) and low (cellular edema) MD lesions, respectively (SM: Table 8 and Table 9). The reliability between manual and automated procedures was high (ICC=0.92) (**Fig. 6**). There was no difference between rater consensus and AQP regarding the determination of brain lesion volumes (P=0.91).

**Figure 5.**
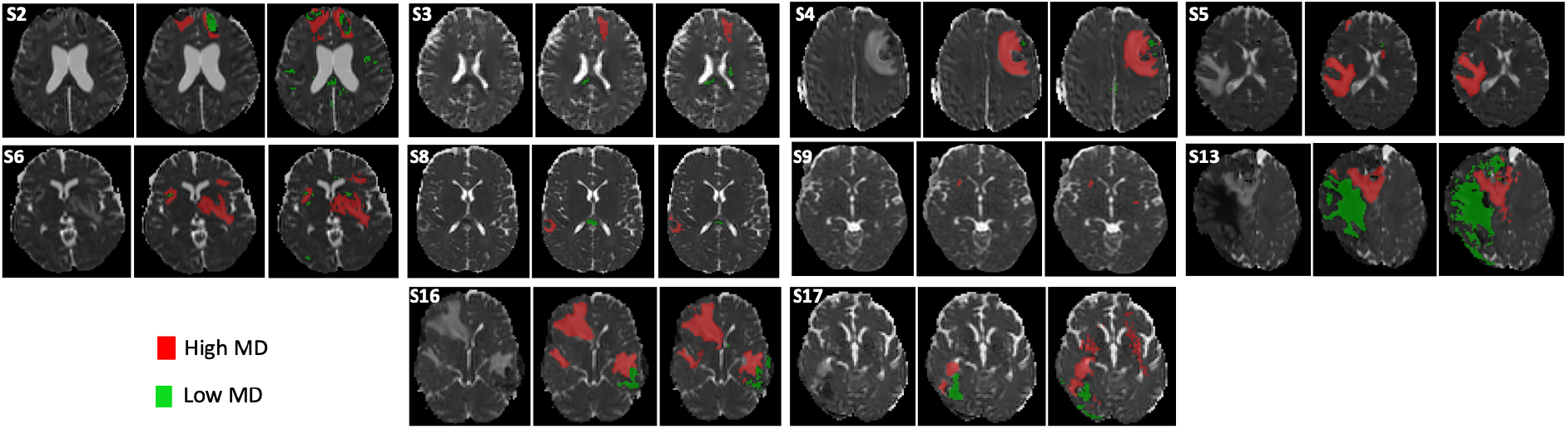
Delineation of brain lesions from diffusion-weighted images (DWI) in 10 TBI patients. For each patient, MD map (left), rater consensus (middle) and automated quantification procedure (right). S2 to S17 refer to the corresponding TBI subject (see Table 3).

**Figure 6.**
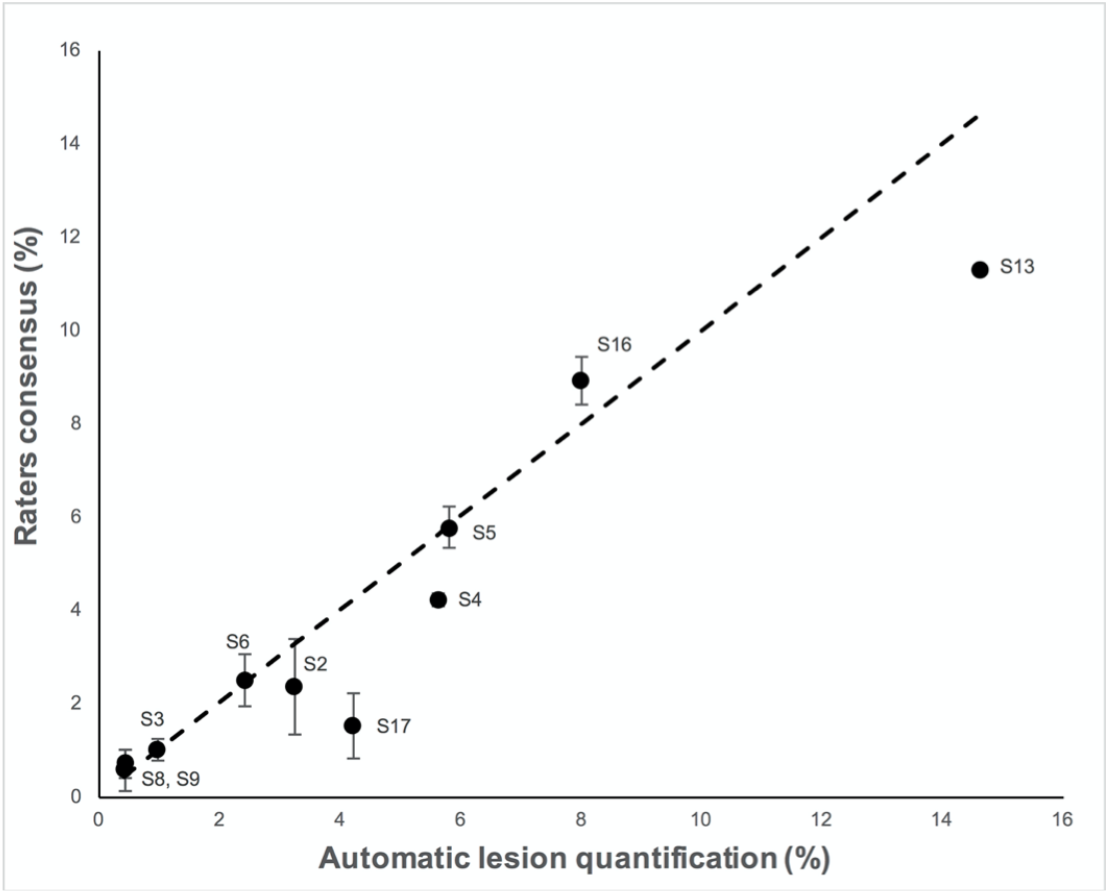
Total lesion volume (low + high MD) in % of the brain volume of diffusion-weighted images (DWI) for ten TBI patients. Lesion volumes (mean, 95% confidence interval) were determined using manual delineation by 5 raters (raters consensus, Y-axis) versus automated quantification procedure (X-axis). The dashed line indicates the identity curve.

For TBI patients, the ICC was higher between AQP and the rater consensus for high MD (0.97) than for low MD (0.48). Similarly, the inter-rater variability was smaller for high (6%) than for low (17%) MD.

## Discussion

Our fully automated procedure (AQP) provided findings in good agreement with manually-traced edematous brain lesions post-trauma. In both realistic phantoms and in TBI patients, both AQP and the expert rater consensus provided comparable lesion volumes with abnormal MD values.

Few studies have explored a fully automated approach to delineate TBI brain lesions. Segmentation method such as Siena, applied to T1-weighted images, misclassified focal TBI lesion in grey matter [21]. Using a deep learning approach, Kamnitsas et al. found 0.63 and 0.68 for Dice and precision, respectively [22]. Better results were obtained using a modified version of the Inception architecture [23], better results were obtained [24]. Our approach permitted quantification of cellular and vasogenic volumes. This approach did not require a training phase with a large set of manual annotations, as required for deep learning approaches. The training phase in our approach is solely based on establishing normal MD distributions in each center for healthy volunteers.

It is interesting to understand the differences between automatic AQP and rater delineation. As seen in Fig 3 and 5, additional brain lesions were found using AQP. Moreover, the contours of the manually-traced ROI were smoother and less detailed than those of the AQP. While these differences had negligible impact on the estimated brain lesion volumes and on the spatial overlap measures (Dice), they can explain the differences in HD, a measure of the maximum distance between two surface points.

**Figure 3.**
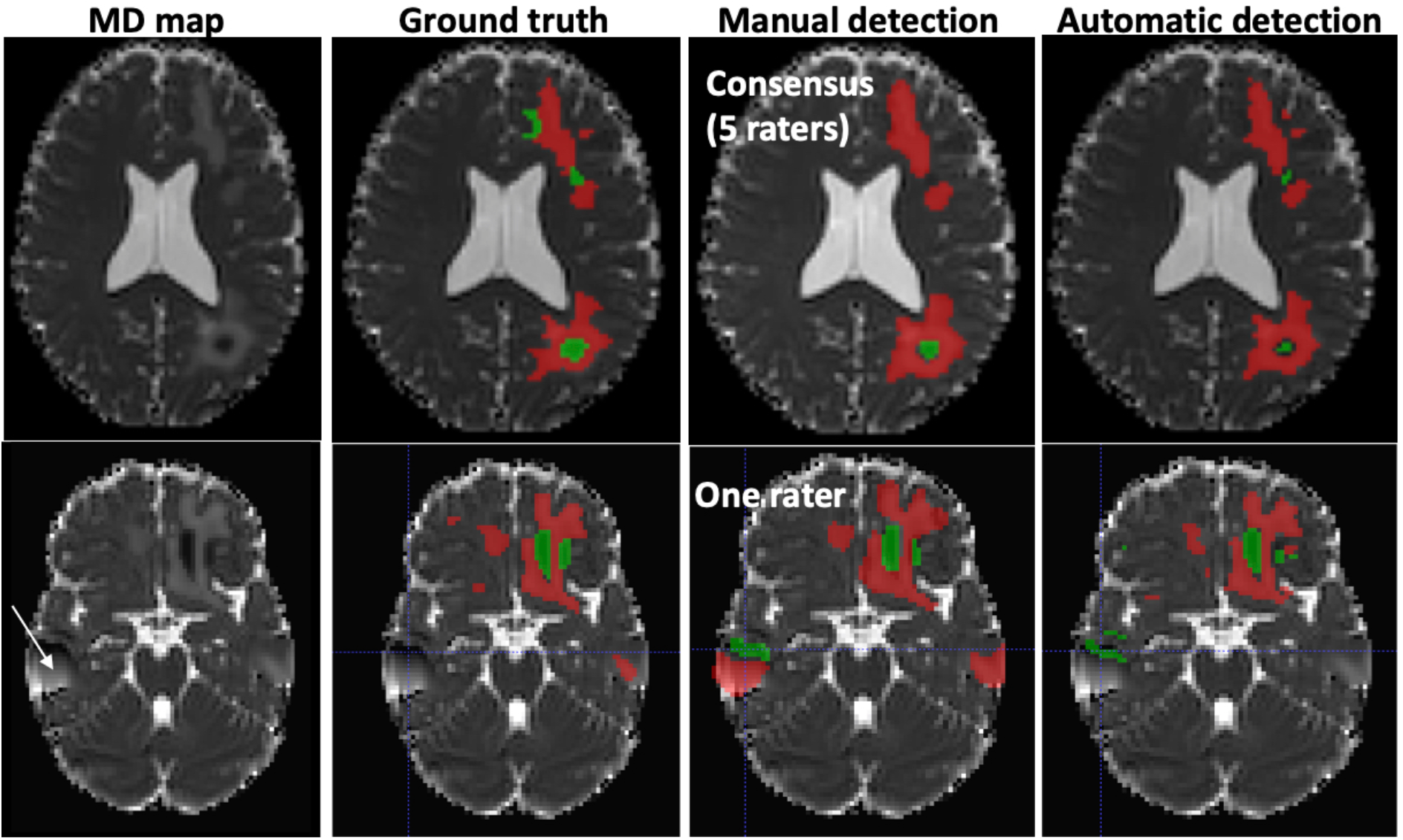
Typical examples of abnormal mean diffusivity (MD) values introduced in diffusion-weighted images (DWI) of two healthy volunteers (realistic TBI phantoms). Top: Good agreement between manual and automatic segmentation. Bottom: Moderate agreement between manual and automatic segmentation. The artefact (white arrow) was falsely detected as a lesion by one rater. Red: High MD values. Green: Low MD values.

Regarding lesion volumes, each manually-traced lesion volume was overestimated (33% in average for phantoms) compared to AQP (5% in average). A closer look at the data shows that the manual delineation systematically overestimated the volume of high MD lesion (44% vs 7% for AQP in average), leading to a smaller ICC between AQP and raters’ consensus for high MD (0.49) than for low MD (0.99).

We observe that Dice similarity coefficient and precision for automated and manual methods were quite low (between 0.59 and 0.70) compared to values obtained for other brain lesion such as stroke or tumor. These low values are indicative of a difficult task, even for experts. This may also explain that the variability of the manual delineation was high (16% for phantoms and 12% for patients).

Limited MRI data exists on the nature of brain injury in the acute phase after severe TBI [5; 7; 25]. Pasco et al. explored the nature of post-traumatic brain edema in the white matter using manual delineation of ROIs based on apparent diffusion coefficient (ADC) values [5]. In the present study, we confirm that both types of brain edema could be found at the early phase of severe TBI. This distinction is not trivial in terms of clinical management: a predominance of lesions with cellular edema (low MD), that reflect brain ischemia, might be required the maintenance of high levels of cerebral perfusion pressure (CCP, with CPP = mean arterial blood pressure – intracranial pressure). On the other hand, lower levels of CPP might be chosen if vasogenic edema (high MD), due to a disruption of the blood-brain barrier, is predominant.

It is important to note that the study of TBI patient management, and associated imaging support, is inherently challenging. As such, the authors draw attention t*o* several limitations. First, brain lesions of realistic TBI phantoms were inserted in brain MD maps only. The use of TBI phantoms with multiparametric images might have resulted in a better agreement with GT. Second, normative MD values were obtained using a limited sample of only 3 young male volunteers per site and TBI data from one patient per site. Although sources of variability between patients and volunteers should be considered, the reliability between manual and automated procedures was however high for TBI patients. Moreover, the number of TBI patients was deliberately limited in this validation study in order to preserve the enrollment of a large number of patients in the OxyTC clinical trial. Third, we considered only one type of MR sequence, i.e. diffusion, and only one metric (MD) for detecting the presence of vasogenic and cellular edema. We did not consider hemorrhagic brain lesions such as contusions, subdural and extradural hematomas, subarachnoid hemorrhage and petechiae, although some may have appeared as low MD lesions. Fourth, the approach seems robust to artefact (see Figure 3). However, the possible misinterpretation of artefacts as lesions requires further studies. Fifth, a larger panel of experts could offer more statistical weight to the results, although it should be noted that we employed the largest panel (5) of experts in TBI imaging, according to the literature [10; 11]. Sixth, a more comprehensive patient dataset to correlate the volume of brain lesions in TBI patients with their outcome was not available.

In conclusion, an automated atlas-based quantification procedure has been effectively shown to quantify the volume of low and high MD brain lesions after trauma, and allow the determination of the nature and volume of edematous brain lesions. This approach had comparable performance with manual delineation by a panel of experts. Even if the involvement of an expert is still necessary to control image quality and validate automatic segmentation, the proposed approach is promising. Indeed, determining the nature and volume of brain edema post-trauma using an accurate and automated approach could support the management of severe TBI patients by directing precision-medicine based treatment for optimal cerebral blood flow. Presently, in a multicenter trial we explore two strategies of patient care management after severe TBI (OxyTC, NCT02754063) using the AQP method.

## Data Availability

MR data supporting the results of this study are available from the corresponding author, on a
collaborative basis.

## Acknowledgements

CM was recipient of a grant from the Gueules Cassées foundation. The authors would like to thank the French network REMI for its assistance in the homogenization of the MR acquisition protocols between imaging centers (France Life Imaging, grants “C7H-FLI11B23” and “C7H-FLI11B19”). Grenoble MRI facility IRMaGe was partly funded by the French program Investissement d’avenir run by the Agence Nationale de la Recherche: grant Infrastructure d’avenir en Biologie Santé ANR-11-INBS-0006.

## Appendix A. Supplementary data

## Abbreviations

(ASSD): Average Symmetrical Surface Distance
AQP: Automatic Quantification Procedure
DWI: Diffusion Weighted Imaging
GT: Ground Truth
(HD): Hausdorff Distance
(ICC): ntra-class Correlation Coefficient
MD: Mean Diffusivity
SM: Supplementary Material
TBI: Train Brain injury

## References

1 Thornhill S, Teasdale GM, Murray GD, McEwen J, Roy CW, Penny KI (2000) Disability in young people and adults one year after head injury: prospective cohort study. BMJ 320:1631–1635

2 Shenton ME, Hamoda HM, Schneiderman JS et al (2012) A review of magnetic resonance imaging and diffusion tensor imaging findings in mild traumatic brain injury. Brain Imaging Behav 6:137–192

3 Kasahara K, Hashimoto K, Abo M, Senoo A (2012) Voxel-and atlas-based analysis of diffusion tensor imaging may reveal focal axonal injuries in mild traumatic brain injury -- comparison with diffuse axonal injury. Magn Reson Imaging 30:496–505

4 Narayana PA, Yu X, Hasan KM et al (2015) Multi-modal MRI of mild traumatic brain injury. Neuroimage Clin 7:87–97

5 Pasco A, Ter Minassian A, Chapon C et al (2006) Dynamics of cerebral edema and the apparent diffusion coefficient of water changes in patients with severe traumatic brain injury. A prospective MRI study. Eur Radiol 16:1501–1508

6 Jha RM, Kochanek PM, Simard JM (2019) Pathophysiology and treatment of cerebral edema in traumatic brain injury. Neuropharmacology 145:230–246

7 Galanaud D, Perlbarg V, Gupta R et al (2012) Assessment of White Matter Injury and Outcome in Severe Brain Trauma A Prospective Multicenter Cohort. Anesthesiology 117:1300–1310

8 Eierud C, Craddock RC, Fletcher S et al (2014) Neuroimaging after mild traumatic brain injury: review and meta-analysis. Neuroimage Clin 4:283–294

9 Wintermark M, Sanelli PC, Anzai Y, Tsiouris AJ, Whitlow CT (2015) Imaging evidence and recommendations for traumatic brain injury: conventional neuroimaging techniques. J Am Coll Radiol 12:e1–14

10 Patel A, van Ginneken B, Meijer FJA, van Dijk EJ, Prokop M, Manniesing R (2017) Robust cranial cavity segmentation in CT and CT perfusion images of trauma and suspected stroke patients. Medical Image Analysis 36:216–228

11 Jain S, Vyvere TV, Terzopoulos V et al (2019) Automatic Quantification of Computed Tomography Features in Acute Traumatic Brain Injury. J Neurotrauma 36:1794–1803

12 Shenton ME, Hamoda HM, Schneiderman JS, et al. (2012) A review of magnetic resonance imaging and diffusion tensor imaging findings in mild traumatic brain injury. Brain Imaging Behav 6:137–192

13 Jones DK, Cercignani M (2010) Twenty-five pitfalls in the analysis of diffusion MRI data. NMR Biomed 23:803–820

14 Shinohara RT, Oh J, Nair G et al (2017) Volumetric Analysis from a Harmonized Multisite Brain MRI Study of a Single Subject with Multiple Sclerosis. AJNR Am J Neuroradiol. 10.3174/ajnr.A5254

15 Chun KA, Manley GT, Stiver SI et al (2010) Interobserver variability in the assessment of CT imaging features of traumatic brain injury. J Neurotrauma 27:325–330

16 Warfield SK, Zou KH, Wells WM (2004) Simultaneous truth and performance level estimation (STAPLE): an algorithm for the validation of image segmentation. IEEE Trans Med Imaging 23:903–921

17 Manjon JV, Coupe P, Concha L, Buades A, Collins DL, Robles M (2013) Diffusion weighted image denoising using overcomplete local PCA. PLoS One 8:e73021

18 Doyle S, Forbes F, Dojat M (2012) P-LOCUS, a complete suite for brain scan segmentation9h IEEE International Symposium on Biomedical Imaging (ISBI),

19 Maggia C, Mistral T, Doyle S et al (2018) Traumatic Brain Lesion Quantification Based on Mean Diffusivity ChangesBrainlesion: Glioma, Multiple Sclerosis, Stroke and Traumatic Brain Injuries, Brainles 2017. (Lecture Notes in Computer Science). Springer-Verlag (Berlin), pp 88–99

20 Watts R, Thomas A, Filippi CG, Nickerson JP, Freeman K (2014) Potholes and molehills: bias in the diagnostic performance of diffusion-tensor imaging in concussion. Radiology 272:217–223

21 Sidaros A, Skimminge A, Liptrot MG et al (2009) Long-term global and regional brain volume changes following severe traumatic brain injury: a longitudinal study with clinical correlates. Neuroimage 44:1–8

22 Kamnitsas K, Ledig C, Newcombe VFJ et al (2017) Efficient multi-scale 3D CNN with fully connected CRF for accurate brain lesion segmentation. Med Image Anal 36:61–78

23 Szegedy C, Liu W, Jia Y et al (2014) Going Deeper with ConvolutionsComputer Vision and Pattern Recognition. arXiv:1409.4842

24 Roy S, Butman JA, Chan L, Pham DL Ieee (2018) TBI Contusion Segmentation from MRI using Convolutional Neural Networks2018 Ieee 15th International Symposium on Biomedical Imaging. (IEEE International Symposium on Biomedical Imaging). arXiv:1807.10839 pp 158–162

25 Ledig C, Heckemann RA, Hammers A et al (2015) Robust whole-brain segmentation: application to traumatic brain injury. Med Image Anal 21:40–58

