## Supplementary material for "Automatic quantification of brain lesion volume from post-trauma MR Images": SM

Supplementary methods – Details of AQP

During the Automated quantification procedure (AQP) (**Fig. 2**), diffusion source images were denoised [1] and hypo or hyper-intense slices were automatically detected. Corrupted slices were corrected by interpolating adjacent slices. Diffusion source images were corrected for inter-volume subject motion and geometric distortion due to susceptibility and eddy current using FMRIB Software Library (FSL) (http://www.fmrib.ox.ac.uk/fsl/) and combined to form diffusion-weighted images. Then, MD maps were computed from the trace of the diffusion-weighted tensors (30 directions, see Table 3). Brain extraction was performed by the ROBEX software (https://sites.google.com/site/jeiglesias/ROBEX) using FLAIR, T2* and T1-weighted images. After registration to the corresponding diffusion-weighted image, brain tissue and CSF were segmented using a Bayesian Markovian approach named PLOCUS [2]. The Montreal Neurological Institute atlas, i.e. MNI152 standard-space T1-weighted average structural template image, was used as *a priori* probabilistic knowledge of tissue localization. Combined with the brain mask provided by ROBEX, this atlas permitted to account for the post-TBI skull deformation. The total brain volume corresponded to the supra-tentorial volume computation.

The AQP used a multi-atlas approach to detect voxels whose values deviated from normative values, according to the Pothole and Molehills method [3]. This multi-atlas approach was based on four atlases: Neuromorphometrics atlas (http://www.neuromorphometrics.com/), HarvardOxford atlas (FSL), Desikan atlas (FreeSurfer), and ICBM DTI81 atlas, to allow the production of 6 brain parcellations corresponding to 1402 regions of interest (ROIs). Using these parcellations and the MRI data of volunteers, a normative MD value was computed for each ROI. For TBI realistic phantoms and TBI patients, the MD value of each voxel was compared to the 6 normative MD values. Abnormal MD values were considered where MD was below the 2$^{nd}$ percentile of normative MD values or above the 94.8$^{th}$ percentile. The asymmetry in thresholds is due to the skewness of the MD distribution. Calculation was repeated for each of the 6 parcellations. A voxel was finally considered as abnormal if 4/6 parcellations at least were found outside the normal range. Thereafter, brain lesion was considered where 20 (or 15) contiguous abnormal voxels at least were observed with high (or low) MD values, respectively, corresponding to a minimal volume of 0.160 (0.120) ml, respectively. Voxels labeled as CSF or ventricles were excluded. To deal with partial volume effect, abnormal voxels at a distance less than 3 mm from CSF voxels were excluded as well. Lesion volume was expressed either in ml or in brain volume fraction (%) that reflects the ratio between brain lesion volume and supra-tentorial brain volume.

1 Manjon JV, Coupe P, Concha L, Buades A, Collins DL, Robles M (2013) Diffusion weighted image denoising using overcomplete local PCA. PLoS One 8:e73021
2 Doyle S, Forbes F, Dojat M (2012) P-LOCUS, a complete suite for brain scan segmentation9h IEEE International Symposium on Biomedical Imaging (ISBI),
3 Watts R, Thomas A, Filippi CG, Nickerson JP, Freeman K (2014) Potholes and molehills: bias in the diagnostic performance of diffusion-tensor imaging in concussion. Radiology 272:217-223

**SM Table 1**: Inclusion and non-inclusion criteria for Patients

**Inclusion Criteria**

- Age between 18 and 75
- Severe non penetrating TBI (initial GCS 3-8) with motor GCS between 1 and 4
- Possible associated extracranial lesions, except tetraplegia
- Monitoring within the first 16 hours since primary traumatic injury
- Indication for ICP monitoring on admission as part of the management
- Indication for continuous sedation/analgesia for more than 48 hours
- Under mechanical ventilation with stable conditions
- Affiliation to the French Social Security or affiliated to a social security system of EU member state, Norway, Lichtenstein,Iceland or Switzerland.
- French-speaking patient

**No inclusion criteria**

- Penetrating TBI
- GCS 3 with bilateral fixed dilated pupils
- Decompressive craniectomy prior to enrolment
- Contraindication of ICP and/or PbtO2 monitoring
- Persistent hemodynamic or respiratory instability
- Hypothermia <34°C on admission
- Venous or arterial lactate concentration >5 mmol/l at randomization
- Life expectancy < 24 hours
- Cardiac arrest at initial presentation
- Tetraplegia
- Neuropsychiatric co-morbidities that could interfere with 6 and 12-months evaluation
- Consent refusal
- Pregnancy
- Participation to another therapeutic study with written consent
- Inability to have a 6-months follow-up
- Permanent contraindications to MRI
- Ischemic stroke after carotid artery dissection
- Incapacitated patients in accordance with article L 1121-5 to L1121-8 of the public health code.

**SM Table 2**: Inclusion and non-inclusion criteria for Controls

**Inclusion criteria**:

- Man between 18 and 60
- No history of chronic disease or brain trauma
- Written informed consent
- Affiliated to the French social security system

**Non-inclusion criteria**:

- Subjects protected by the law (art. L1121-6 and L1121-8)
- Contra-indications to MRI
- Claustrophobia

**SM Table 3**: Main MR acquisition parameters for the different scanners IT. NSA: Number of acquisitions. Na: Not available.

| 3T | 3D FLAIR | | 3D T1 MPRAGE | | T2* | | DWI | | b0 | |
|---|---|---|---|---|---|---|---|---|---|---|
| | Philips | Siemens | Philips | Siemens | Philips | Siemens | Philips | Siemens | Philips | Siemens |
| Orientation | sagittal | | sagittal | | transversal | | transversal | | transversal | |
| TR / TE (ms) | 4800/390 | 5000/402 | 2500/3.3 | 2300/2 | 2200/16 | 1110/12 | 9800/80 | 10900/80 | 9800/80 | 10900/80 |
| TI (ms) | 1650 | 1800 | 940 | 900 | | | | | | |
| Turbo / EPI | 182 | 270 | 232 | 208 | | | Single shot EPI | | Single shot EPI | |
| Resolution (mm$^3$) | 1x1x1 | | 1x1x1 | | 1x1x2 | | 2x2x2 | | 2x2x2 | |
| Flip angle | 90° | 120° variable | 9° | | 16° | 20° | 90° | | 90° | |
| Slices | 180 | 176 | 180 | 176 | 70 | 66 | 70 | | 70 | |
| Gap (mm) | 0 | | 0 | | 0 | | 0 | | 0 | |
| NSA | 2 | 1 | 1 | | 1 | | 1 | | 1 | |
| Diffusion direction | | | | | | | 1 b=0 32 b=1000 | 1 b=0 30 b=1000 | 1 b=0 | |
| Phase direction | | | | | | | P >> A | | A >> P | |

| 1.5T | 3D FLAIR | | 3D T1 MPRAGE | | T2* | | DWI | | b0 | |
|---|---|---|---|---|---|---|---|---|---|---|
| | GE | Siemens | GE | Siemens | GE | Siemens | GE | Siemens | GE | Siemens |
| Orientation | sagittal | | sagittal | | transversal | | transversal | | transversal | |
| TR / TE (ms) | 8000/180 | 5000/335 | 6.2/2 | 2000/2.9 | 720/23 | 1230/20 | 12000/85 | 6700/85 | 12000/85 | 6700/85 |
| TI (ms) | 2100 | 1800 | 450 | 1100 | | | | | | |
| Turbo / EPI | 213 | 242 | ? | 208 | | | Single shot EPI | | Single shot EPI | |
| Resolution (mm$^3$) | 1x1x1 | | 1x1x1 | | 1x1x3 | | 2.5x2.5x2.5 | | 2.5x2.5x2.5 | |
| Flip angle | Na | 120° | 12° | 15° | 20° | | 90° | | 90° | |
| Slices | 156 | 176 | 160 | 160-192 | 48 | 44 | 55 | | 55 | |
| Gap (mm) | Na | 0 | 0 | | 0 | | 0 | | 0 | |
| NSA | 2 | 1 | 1 | | 2 | 1 | 1 | | 1 | |
| Diffusion directions | | | | | | | 1 b=0 30 b=1000 | | 1 b=0 30 b=1000 | |
| Phase direction | | | | | | | P >> A | | A >> P | |

**SM Table 4**: Volume, expressed in % of the brain volume, of the lesion for each realistic TBI phantoms for the automated method, each rater and staple computed on the five raters scores.

Difference in lesion volume are expressed in % of the GT (Ground Truth) lesion. AQP: automatic quantification procedure.

| Case | GT Vol (%) | AQP | Rater 1 | Rater 2 | Rater 3 | Rater 4 | Rater 5 | Rater consensus | AQP-GT (%) | Rater consensus-GT (%) |
|---|---|---|---|---|---|---|---|---|---|---|
| 1 | 1.1 | 1.2 | 1.4 | 1.0 | 1.1 | 1.03 | 1.7 | 1.4 | 11.0 | 24.8 |
| 2 | 1.6 | 1.8 | 2.2 | 1.5 | 1.9 | 1.8 | 1.9 | 2.1 | 10.6 | 32.3 |
| 3 | 2.1 | 2.6 | 2.5 | 2.0 | 2.5 | 2.2 | 2.9 | 2.7 | 25.2 | 33.5 |
| 4 | 2.4 | 1.7 | 3.2 | 2.5 | 2.6 | 2.0 | 3.5 | 3.3 | -30.7 | 35.7 |
| 5 | 3.6 | 3.8 | 4.1 | 3.6 | 3.8 | 4.4 | 5.0 | 4.8 | 7.2 | 35.1 |

**SM Table 5**: Volume expressed in ml of the high MD lesion for each realistic TBI phantoms for the automatic quantification procedure (AQP), each rater and staple computed on the five raters scores. Difference in lesion volume are expressed in % of the GT (Ground Truth) lesion.

| Case | GT (ml) | AQP (ml) | Rater 1 (ml) | Rater 2 (ml) | Rater 3 (ml) | Rater 4 (ml) | Rater 5 (ml) | Rater consensus (ml) | AQP-GT (%) | Rater consensus-GT (%) |
|---|---|---|---|---|---|---|---|---|---|---|
| 1 | 24.2 | 26.2 | 34.2 | 25.1 | 29.3 | 27.7 | 29.8 | 33.9 | 9.0 | 41.0 |
| 2 | 16.2 | 16.7 | 22.3 | 16.6 | 17.3 | 15.9 | 27.3 | 21.7 | 5.0 | 34.3 |
| 3 | 31.2 | 38.0 | 39.4 | 30.3 | 38.7 | 32.4 | 44.3 | 42.1 | 21.82 | 35.1 |
| 4 | 32.9 | 22.0 | 46.8 | 36.0 | 39.5 | 29.8 | 51.2 | 48.8 | -33.2 | 48.3 |
| 5 | 24.9 | 32.8 | 34.8 | 29.9 | 31.5 | 37.6 | 39.6 | 39.5 | 31.84 | 58.9 |

**SM Table 6**: Volume expressed in ml of the low MD lesion for each realistic TBI phantoms for the automatic quantification procedure (AQP). each rater and staple computed on the five raters scores. Difference in lesion volume are expressed in % of the GT (Ground Truth) lesion.

| Case | GT (ml) | AQP (ml) | Rater 1 (ml) | Rater 2 (ml) | Rater 3 (ml) | Rater 4 (ml) | Rater 5 (ml) | Rater consensus (ml) | AQP-GT (%) | Rater consensus-GT (%) |
|---|---|---|---|---|---|---|---|---|---|---|
| 1 | 3.5 | 4.4 | 2.9 | 1.2 | 2.5 | 2.6 | 2.8 | 2.8 | 26.0 | -27.7 |
| 2 | 1.7 | 2.9 | 1.1 | 0.5 | 0.6 | 1.0 | 0.4 | 0.4 | 72.0 | -60.7 |
| 3 | 2.9 | 4.8 | 2.8 | 2.7 | 3.2 | 3.3 | 3.1 | 3.1 | 63.7 | 17.1 |
| 4 | 4.0 | 3.6 | 1.1 | 1.5 | 0 | 1.2 | 2.5 | 2.5 | -10.8 | -69.0 |
| 5 | 15.0 | 9.8 | 11.2 | 10.0 | 11.1 | 11.5 | 15.5 | 15.5 | -34.3 | -4.7 |

**SM Table 7**: All mean spatial measures for the 10 TBI patients for each rater. For each rater, the ground truth (GT) was computed using the Staple method on the results of the two other raters.

Median [25th, 75th].

| Comparison | Dice | Hausdorff distance (mm) | ASSD (mm) | Precision | Sensitivity |
|---|---|---|---|---|---|
| Rater 1 vs GT | 0.56 [0.46 0.74] | 20.4 [15.6 27.6] | 1.5 [0.9 2.2] | 0.42 [0.32 0.69] | 0.82 [0.75 0.89] |
| Rater 2 vs GT | 0.58 [0.40 0.67] | 21.6 [16.5 26.5] | 1.3 [1.0 2.1] | 0.44 [0.27 0.59] | 0.84 [0.80 0.89] |
| Rater 3 vs GT | 0.67 [0.54 0.74] | 16.6 [9.1 19.6] | 1.14 [0.7 1.8] | 0.67 [0.65 0.75] | 0.70 [0.58 0.76] |
| Median | 0.59 [0.44 0.74] | 19.6 [13.6 26.1] | 1.4 [0.8 2.2] | 0.60 [0.33 0.71] | 0.79 [0.70 0.88] |

**SM Table 8**: Volume expressed in ml of the high MD lesion (vasogenic edema) for each patient for the automatic quantification procedure (ACQ), each rater and staple computed on the three raters scores (** two raters, * 1 rater). Difference in lesion volume are expressed in % of the consensus lesion volume. Na: not available.

| Subject | AQP (ml) | Rater 1 (ml) | Rater 2 (ml) | Rater 3 (ml) | Rater consensus (ml) | AQP – Rater consensus (%) |
|---|---|---|---|---|---|---|
| 2 | 36.8 | 37.5 | 23.0 | 33.2 | 26.8 | 37.4 |
| 3 | 8.7 | 14.6 | 10.4 | 12.2 | 11.4 | -23.7 |
| 4 | 67.8 | 79.2 | Na | 69.0 | 57.7** | 17.5 |
| 5 | 79.2 | 89.0 | 79.9 | 85.6 | 82.2 | -3.6 |
| 6 | 28.4 | 30.4 | 27.2 | 37.5 | 28.8 | -1.5 |
| 8 | 5.7 | 16.7 | 10.8 | 14.0 | 9.8 | -41.6 |
| 9 | 3.0 | 6.3 | 0.621 | 7.1 | 2.0 | 50.8 |
| 13 | 72.9 | - | 74.1 | - | 74.1* | -1.6 |
| 16 | 121.0 | 111.4 | 113.9 | - | 94.5** | 28,1 |
| 17 | 45.1 | 10.0 | 10.7 | - | 7.15** | 530,6 |

**SM Table 9**: Volume expressed in ml of the low MD lesion (cellular edema) for each patient for the automatic quantification procedure (ACQ), each rater and staple computed on the three raters scores (** two raters, * 1 rater). Difference in lesion volume are expressed in % of the consensus lesion volume. Na: not available.

| Subject | AQP (ml) | Rater 1 (ml) | Rater 2 (ml) | Rater 3 (ml) | Rater consensus (ml) | AQP – Rater consensus (%) |
|---|---|---|---|---|---|---|
| 2 | 18.4 | 16.7 | 1.1 | 20.7 | 10.0 | 83.0 |
| 3 | 5.2 | 5.0 | 2.7 | 5.5 | 2.6 | 102.4 |
| 4 | 13.1 | 3.3 | Na | 11.0 | 1.6** | 699.0 |
| 5 | 5.3 | 2.5 | 0 | 4.8 | 1.4 | 288.6 |
| 6 | 4.7 | 5.3 | 0 | 8.7 | 4.4 | 7.1 |
| 8 | 1.5 | 2.1 | 0.6 | 6.1 | 1.0 | 44.4 |
| 9 | 3.9 | 6.0 | 5.6 | 13.1 | 6.3 | -38.0 |
| 13 | 167.7 | nan | 111.5 | nan | 111.5* | 50.4 |
| 16 | 9.2 | 14.3 | 0 | nan | 14.3** | -35.3 |
| 17 | 18.4 | 12.4 | 0 | nan | 12.4** | 48.6 |
